# Hydroxychloroquine with or without azithromycin and in-hospital mortality or discharge in patients hospitalized for COVID-19 infection: a cohort study of 4,642 in-patients in France

**DOI:** 10.1101/2020.06.16.20132597

**Authors:** Emilie Sbidian, Julie Josse, Guillaume Lemaitre, Imke Meyer, Mélodie Bernaux, Alexandre Gramfort, Nathanaël Lapidus, Nicolas Paris, Antoine Neuraz, Ivan Lerner, Nicolas Garcelon, Bastien Rance, Olivier Grisel, Thomas Moreau, Ali Bellamine, Pierre Wolkenstein, Gaël Varoquaux, Eric Caumes, Marc Lavielle, Armand Mekontso Dessap, Etienne Audureau, On behalf of AP-HP/Universities/Inserm COVID-19 research collaboration, AP-HP Covid CDR Initiative and ‘Entrepôt de Données de Santé’ AP-HP consortium

**Affiliations:** University Paris Est Creteil, Reserach unit EpiDermE, Creteil, France; AP-HP, Henri-Mondor hospital, Department of Dermatology, Creteil, France; INSERM, Centre d’Investigation Clinique 1430, Créteil, France; Department of Statistics INRIA, Saclay Paris Sud University, France; Ecole Polytechnique, Palaiseau, France; Parietal, Inria Saclay, Palaiseau, France; EHESS (Ecole des Hautes études en sciences sociales); Strategy and transformation department, APHP Greater Paris University Hospital; Sorbonne Université, INSERM, Institut Pierre Louis d’Epidémiologie et de Santé Publique IPLESP, AP-HP. Sorbonne Université, Public Health Department, Saint-Antoine Hospital, Paris, France; WIND Department APHP Greater Paris University Hospital; INSERM, UMRS 1138, Cordeliers research center, Sorbonne Université, Paris, France; Département d’informatique médicale, Hôpital Necker-Enfants Malades, Assistance Publique - Hôpitaux de Paris (AP-HP), F-75015 Paris, France; Institut Imagine, Université Paris Descartes - Université de Paris, F-75015 Paris, France; Département d’informatique médicale, Hôpital Européen Georges Pompidou, Assistance Publique - Hôpitaux de Paris (AP-HP), F-75015 Paris, France; APHP, Hôpitaux Universitaires Pitié Salpêtrière-Charles Foix, Infectious Diseases department, Paris, France; Sorbonne Université, Institut National de la Santé et de la Recherche Médicale (INSERM), Institut Pierre-Louis d’Epidémiologie et de Santé Publique, Paris, France; Medical Intensive Care Unit, Henri Mondor Hospital, Créteil, France; Inserm U955, Université Paris Est Créteil (UPEC), Institut Mondor de Recherche Biomédicale (IMRB), équipe CEpiA (Clinical Epidemiology and Ageing), Créteil, France; AP-HP, Henri-Mondor hospital, Department of Public Health, Clinical Research Unit, Creteil, France

**Keywords:** Covid-19 infection, Hydroxychloroquine, Death, hospital discharge

## Abstract

**Objective:** To assess the clinical effectiveness of oral hydroxychloroquine (HCQ) with or without azithromycin (AZI) in preventing death or leading to hospital discharge.

**Design:** Retrospective cohort study.

**Setting:** An analysis of data from electronic medical records and administrative claim data from the French Assistance Publique - Hôpitaux de Paris (AP-HP) data warehouse, in 39 public hospitals, Ile-de-France, France.

**Participants:** All adult inpatients with at least one PCR-documented SARS-CoV-2 RNA from a nasopharyngeal sample between February 1^st^, 2020 and April 6^th^, 2020 were eligible for analysis. The study population was restricted to patients who did not receive COVID-19 treatments assessed in ongoing trials, including antivirals and immunosuppressive drugs. End of follow-up was defined as the date of death, discharge home, day 28 after admission, whichever occurred first, or administrative censoring on May 4, 2020.

**Intervention:** Patients were further classified into 3 groups: (i) receiving HCQ alone, (ii) receiving HCQ together with AZI, and (iii) receiving neither HCQ nor AZI. Exposure to a HCQ/AZI combination was defined as a simultaneous prescription of the 2 treatments (more or less one day).

**Main outcome measures:** The primary outcome was all-cause 28-day mortality as a time-to-event endpoint under a competing risks survival analysis framework. The secondary outcome was 28-day discharge home. Augmented inverse probability of treatment weighted (AIPTW) estimates of the average treatment effect (ATE) were computed to account for confounding.

**Results:** A total of 4,642 patients (mean age: 66.1 ± 18; males: 2,738 (59%)) were included, of whom 623 (13.4%) received HCQ alone, 227 (5.9%) received HCQ plus AZI, and 3,792 (81.7%) neither drug. Patients receiving ‘HCQ alone’ or ‘HCQ plus AZI’ were more likely younger, males, current smokers and overall presented with slightly more co-morbidities (obesity, diabetes, any chronic pulmonary diseases, liver diseases), while no major difference was apparent in biological parameters. After accounting for confounding, no statistically significant difference was observed between the ‘HCQ’ and ‘Neither drug’ groups for 28-day mortality: AIPTW absolute difference in ATE was +1.24% (−5.63 to 8.12), ratio in ATE 1.05 (0.77 to 1.33). 28-day discharge rates were statistically significantly higher in the ‘HCQ’ group: AIPTW absolute difference in ATE (+11.1% [3.30 to 18.9]), ratio in ATE (1.25 [1.07 to 1.42]). As for the ‘HCQ+AZI’ vs neither drug, trends for significant differences and ratios in AIPTW ATE were found suggesting higher mortality rates in the former group (difference in ATE +9.83% [-0.51 to 20.17], ratio in ATE 1.40 [0.98 to 1.81];p=0.062).

**Conclusions:** Using a large non-selected population of inpatients hospitalized for COVID-19 infection in 39 hospitals in France and robust methodological approaches, we found no evidence for efficacy of HCQ or HCQ combined with AZI on 28-day mortality. Our results suggested a possible excess risk of mortality associated with HCQ combined with AZI, but not with HCQ alone. Significantly higher rates of discharge home were observed in patients treated by HCQ, a novel finding warranting further confirmation in replicative studies. Altogether, our findings further support the need to complete currently undergoing randomized clinical trials.

**WHAT THIS PAPER ADDS?:** *What is already known on this subject:*

- The use of Hydroxychloroquine (HCQ) or HCQ with azithromycin (AZI) has been associated with viral load reduction at 6 days in COVID-19 infected patients
- No difference between HCQ and no-HCQ groups in terms of risk of death or need for mechanical ventilation was found in two large cohorts of hospitalized COVID-19 infected patients

*What this study adds:*

- Using a large non-selected population of inpatients hospitalized for COVID-19 infection in 39 hospitals in France and robust methodological approaches, we found no evidence for efficacy of HCQ on 28-day mortality
- Our results suggest an excess risk of mortality in patients treated by a combination of HCQ and AZI, but not with HCQ alone
- Significantly higher rates of discharge home were observed in patients treated by HCQ, a novel finding warranting further confirmation in replicative studies

## Introduction

The COVID-19 pandemic due to the SARS-CoV-2 coronavirus started in Wuhan, China last December, 2019.^1^ The COVID-19 epidemic is a worldwide pandemic with more than 5.4 million cases reported up to May 27, 2020, of whom nearly 350,000 have died. Because effective treatments are urgently needed, more than 600 clinical trials are currently ongoing in a worldwide effort to fight the coronavirus.

The 4-aminoquinolines chloroquine (CQ) and hydroxychloroquine (HCQ) are synthetic antimalarials drugs (AMD). Due to their anti-inflammatory and immunomodulatory properties, synthetic AMDs are the standard treatment in autoimmune diseases such as lupus. In vitro data schowed a non-specific antiviral action of synthetic AMDs against emerging viruses such as HIV, dengue, hepatitis C, chikungunya, influenza, Ebola, severe acute respiratory syndrome, and middle east respiratory syndrome and human immunodeficiency viruses.^2^

The use of HCQ or HCQ with azithromycin (AZI) has arisen as a promising treatment or combination of treatment for COVID-19 infection. First, in vitro data have shown the effectiveness of HCQ (and to a lesser extent CQ) in reducing the viral load of cells infected with SARS-CoV-2.^3^ Then, a Chinese clinical trial showed that CQ had a significant effect, including a better clinical outcome, when compared to control groups.^4^ A French research team suggested that HCQ, at a dose of 600 mg/day, was associated with viral load reduction in twenty COVID-19 patients and its effect was strengthened by AZI.^5^ These preliminary results were further backed up by two prospective cohorts from the same team of 80 and 1,061 participants, suggesting good clinical outcomes in 65 (81%) and 973 (92%) patients, respectively, with lower frequency of aggressive clinical course requiring oxygen therapy, fewer transfers to the intensive care unit (ICU), or death after at least 3 days of treatment and a viral load reduction at day 6.^6,7^

More recently, several published or preprint publications have raised the question of HCQ efficacy for COVID-19 infection. Four observational studies with diverse cohort sizes (81, 368, 1,376, and 1,438 participants) failed to find a difference between HCQ and no-HCQ groups in terms of risk of death or need for mechanical ventilation.^8–11^ Preliminary results from the UK RECOVERY randomized trial have been communicated, but not yet published, concluding that there was no beneficial effect of HCQ on 28-day mortality in hospitalized patients with COVID-19.^12^

Data from large health care databases provide a unique opportunity to assess the potential effectiveness and harm of HCQ in a real-world setting, including unselected population. Because some variation has been reported between studies, replicated analyses minimizing selection and confounding biases are crucially needed to disentangle current evidence on the actual risks and benefits of HCQ-based treatments. We consequently aimed to assess the clinical effectiveness of oral HCQ in preventing death or allowing to hospital discharge using a large, exhaustive, non-selected population of in-patients hospitalized for COVID-19 infection in 39 hospitals in France, accounting in detail for patients characteristics.

## Methods

### Study Design

We performed a retrospective cohort study using the Corona OMOP database, which combines electronic medical records and administrative claim data from the Greater Paris Public Hospitals (Assistance Publique - Hôpitaux de Paris (AP-HP) data warehouse, called ‘Entrepôt de Données de Santé’ (EDS). This study was approved by the French data protection agency Commission Nationale de l’Informatique et des Libertés (regulatory decision DE-2017-013), IRB00011591. Patients and the public were not involved in the present study.

### Data sources

The EDS currently collects data on more than 11 million patients treated in the 39 hospitals of the AP-HP, Ile-de-France, France. The warehouse contains medico-administrative data from the Medical Information System Program (PMSI), the French national hospital database which gathers information from standardized discharge reports on diagnoses and procedures performed in all medical units involved in patient management during his/her hospital stay. Primary and associated diagnoses are recorded using the International Classification of Diseases, 10^th^ edition (ICD-10) and therapeutic procedures using a national standardized classification system (*Classification Commune des Actes Médicaux*, CCAM, 11^th^ edition). Since discharge reports are mandatory and used for hospital fund allocation, the PMSI database contains exhaustive information on all admissions to hospitals in France, including proprietary and public hospitals.

In addition to PMSI data, the EDS gathers information from multiple electronic health record databases, including biology and imaging results, drug prescriptions (stored within the ORBIS medication database and coded according to the international Anatomical Therapeutic Chemical (ATC) classification system) and medical text reports associated with hospital visits, including emergency department data and outpatient visits.

### Data acquisition

Because PMSI coding requires human-based and time-consuming processing, complete PMSI information was not available for a number of patients being still hospitalized or for whom discharge reports were not yet processed at the time of analysis. Likewise, information on drugs prescriptions were not directly available in some ICU not using the ORBIS medication system. Consequently, data acquisition for the present study relied on both structured data (i.e. PMSI pertaining to past hospitalizations, if any, biological results, ORBIS medication system) and unstructured data (i.e. medical text records). For the latter, we used artificial intelligence algorithms based on Natural Language Processing (NLP), to extract information on patients diagnoses (including comorbidities, see below) and drugs prescriptions (including HCQ+/-AZI), considering contexts where mentions of drugs by name do not correspond to actual prescriptions (i.e. when the drug is mentioned in a negative context),^13^ and considering both International non-proprietary name (INN) and trade-marks.

### Study population

All adult (≥ 18 years of age) inpatients with at least one polymerase chain reaction-documented SARS-CoV-2 RNA from a nasopharyngeal sample between February 1^st^, 2020 and April 6^th^, 2020 were eligible for the present analysis. The date of inclusion in the study cohort (index date) was defined as the date of admission. We restricted the study population to previously hydroxychloroquine- and azithromycin-naive inpatients, defined as those who had not received a prescription before the index date, and excluded those having received specific COVID-19 treatments, i.e. treatments assessed in ongoing trials: remdesivir, lopinavir-ritonavir (ATC J05AR10), favipiravir (J05AX27), anti-interleukin 1 - i.e., anakinra (ATC L04AC03), canakinumab (ATC L04AC038) - anti-interleukin 6 - i.e., tocilizumab (ATC L04AC037), sarilumab (ATC L04AC14). When patients were transferred between hospitals for the same stay, several discharge reports were available which were analyzed as a single hospital stay until first discharge home. Patients who died or were discharged within 24 hours following their admission were excluded. The end of follow-up was defined by the time of death, discharge home, day 28 (D28) after admission, whichever occurred first, or administrative censoring on May 4, 2020. Patients transferred to hospitals outside AP-HP or to follow-up care and rehabilitation services before day 28 were considered as censored.

### Outcomes

The study’s primary outcome was all-cause 28-day mortality, assessed as a time-to-event endpoint under a competing risks survival analysis framework. For patients discharged home before day 28, we looked at subsequent re-admissions to determine vital status on day 28. The secondary outcome was 28-day discharge home, also assessed as a time-to-event endpoint.

### Drug exposures

While there was no overarching recommendation regarding HCQ+/-AZI prescription at the AP-HP level, guidelines were nonetheless proposed at local level in several individual hospitals, suggesting hydroxychloroquine to physicians as a therapeutic option for patients with moderate-to-severe COVID-19 infection, i.e. requiring oxygen. The suggested HCQ regimen was a loading dose of 600 mg on day 1, followed by 400 mg daily for 9 additional days. AZI at a dose of 500 mg on day 1 and then 250 mg daily for 4 more days in combination with HCQ was an additional suggested therapeutic option. Prescription of HCQ or HCQ together with AZI was at the discretion of the physicians.

Using data acquisition procedures previously detailed, we identified patients with a prescription of HCQ (ATC P01BA02), AZI (ATC J01FA10), steroids (ATC H02AB), and antithrombotic agents (heparin group, ATC B01AB) usually used for acute respiratory distress syndrome.^14,15^ Exposure to a HCQ/AZI combination was defined as a simultaneous prescription of the two treatments (within one day). Based on previous possible combinations, patients were further classified into three groups: (i) receiving HCQ alone, (ii) receiving HCQ together with AZI, and (iii) receiving neither HCQ nor AZI. Patients receiving AZI alone were excluded, in accordance with our objective to assess clinical effectiveness of HCQ with or without AZI vs. neither specific treatment.

### Covariates

For each patient, age, sex, hospital-admission location, ICU admission, ICU stay length and hospital stay length were recorded. Co-morbidities and risk factors (smoker status, obesity, hypertension, diabetes, dyslipidemia, ischaemic or rhythmic heart diseases, heart failure, renal disease, presence of chronic respiratory insufficiency or asthma or cystic fibrosis, and cancer) were recorded for the two-year period before the index date. Clinical severity features within 24h after admission were also recorded: ICU transfer within the first 24h, oxygen saturation, partial pressure of oxygen (PaO_2_ mmHg), and carbon dioxyde (PCO_2_). Lastly, biologic values were also recorded at the index date using their LOINC (Logical Observation Identifiers Names and Codes). Biological values were recorded for neutrophils, lymphocytes, C reactive protein, D-Dimer, prothrombin time, creatine, and lacticodesydrogenase. Detailed definitions of the covariates are available in **Supplemental Table 1**.

### Statistical analysis

All analyses were performed considering the three main treatment modalities of interest, as previously described (HCQ/HCQ+AZI/Neither drug), in the entire population or after stratifying by the level of severity of COVID-19 at admission, considering (i) early ICU admission as defined as occurring within the first 24 hours of admission or (ii) not.

#### Descriptive statistics

Descriptive results are given as medians [interquartile range (IQR)] for continuous variables, and counts (%) for categorical variables. Unadjusted between-groups comparisons were performed using the Chi-square test for comparing categorical variables, the Kruskal-Wallis (three-groups comparisons) and Mann–Whitney rank sum tests (pairwise comparisons, correcting for test multiplicity with the Benjamini-Hochberg procedure) for continuous variables, as appropriate. For time-to-event analyses, non-parametric Nelson–Aalen estimates of the cumulative cause-specific hazards were plotted for the occurrence of death or hospital discharge.

#### Causal inference modeling

Due to the influence on treatment assignment of baseline characteristics of patients included in non-randomized observational studies, it is essential to account for such differences when estimating treatment effects to address bias arising from confounding.^16^ Under assumptions of unconfoundedness (i.e., potential outcomes are independent of the treatment assignment conditionally on a vector of covariates)^17^ and its extension in the presence of missing values,^18^ the average treatment effect (ATE) can be identified.^19^ For the present analysis, causal inference modeling relied on doubly robust estimators, combining an outcome regression with a model for the treatment (i.e., propensity score) to derive augmented inverse probability of treatment weighting (AIPTW) estimators, a more effective approach in minimizing bias due to model misspecification than only IPTW.^20^

The selection of the relevant covariates to be used in causal inference modeling was based on available published data at the time of analysis^21^ and expert *a priori* knowledge on key prognostic factors and determinants of treatment assignment, including patients’ demographics, co-morbidities, hospital and time period of admission. **Supplemental Figure 1** generated using

DAGitty,^22^ shows the causal inference model we applied, differentiating between variables assessed as predictors of the treatment assignment, unrelated to the outcome (brown), predictors of the outcome, unrelated to the treatment assignment (blue), predictors of both treatment and outcome (violet).

As the primary analysis, we constructed cause-specific Cox proportional hazards regression models to account for the competing risk between all-cause death and hospital discharge. Doubly-robust estimation equations were derived based on regression models for the outcome and the censoring (using Cox modeling), and the treatment distribution (using a generalized linear model with a logit link function), conditional on baseline covariates.^23^ ATEs were calculated as the absolute difference and ratios in the standardized absolute risks,^24^ along with their 95% confidence intervals.

#### Sensitivity analyses and missing data handling

We conducted several sensitivity analyses to check the stability of our results under varying approaches. First, we assessed 28-day mortality as a binary endpoint, considering patients discharged home before day 28 as alive at that date and excluding patients transferred to hospitals outside AP-HP or to follow-up care and rehabilitation services before day 28. To do so, we used the causal forest implementation based on the generalized random forests (GRF) method^25^ to compute forest-based weighting functions and derive AIPTW estimates for doubly robust inference of the average treatment effect (ATE). Second, conventional multivariable and IPT-weighted analyses (‘singly robust’) using cause specific Cox models and Fine-Gray competing risks analyses were also performed, computing IPT-weighted and adjusted hazard ratios (HR) and subhazard ratios (SHR), respectively, and plotting raw and adjusted cumulative incidence curves to illustrate the associations. For all IPT-weighting based analyses, standardized differences of the means were computed before and after IPTW to assess imbalance of the covariates between treatment groups. Standardized differences less than 10% were considered negligible following common practice when using IPTW to estimate causal treatment effects in observational studies.^26^

To account for the potential influence of missing data on causal inference procedures, we used single imputation with a (regularized) iterative Factorial Analysis for Mixed Data model (FAMD), accounting for similarities between both individuals and relationships between covariates, treatment assignment and the outcome. Variables showing departure from normality using graphical methods and kurtosis/skewness statistics were log-transformed prior to imputation. For the GRF method, the GRF-MIA approach which enables the computation of ATE without any imputation of the data was used for missing data handling.

A two-tailed p-value of less than 0.05 was considered significant. Statistical analyses were performed using R v3.6.4 (R Foundation for Statistical Computing, Vienna, Austria; packages *grf, riskRegression*).^23,25^

## Results

### Study population

From February 1^st^ to April 6, 2020, 5,556 adult patients were hospitalized at AP-HP for a Covid-19 infection and did not receive specific COVID-19 treatments other than HCQ or AZI. Patients who died (n=91) or who were discharged (n=196) within 24 hours after their admission were excluded, as well as patients receiving AZI alone (n=582) or patients who did not initiate HCQ and AZI the same day (more or less 24 hours, n=45). Thus, a total of 4,642 patients (mean age: 66.1±18; males: 2,738 (59%)) were included in the study population, of whom 623 (13.4%) received HCQ alone, 227 (5.9%) received HCQ plus AZI, and 3,792 (81.7%) neither HCQ nor HCS plus AZI (**Figure 1**). In the ‘HCQ alone’ and ‘HCQ plus AZI’ groups, median timing of the first dose of HCQ after the admission was 0.42 days, IQR (0 to 2.3).

**Figure 1.**
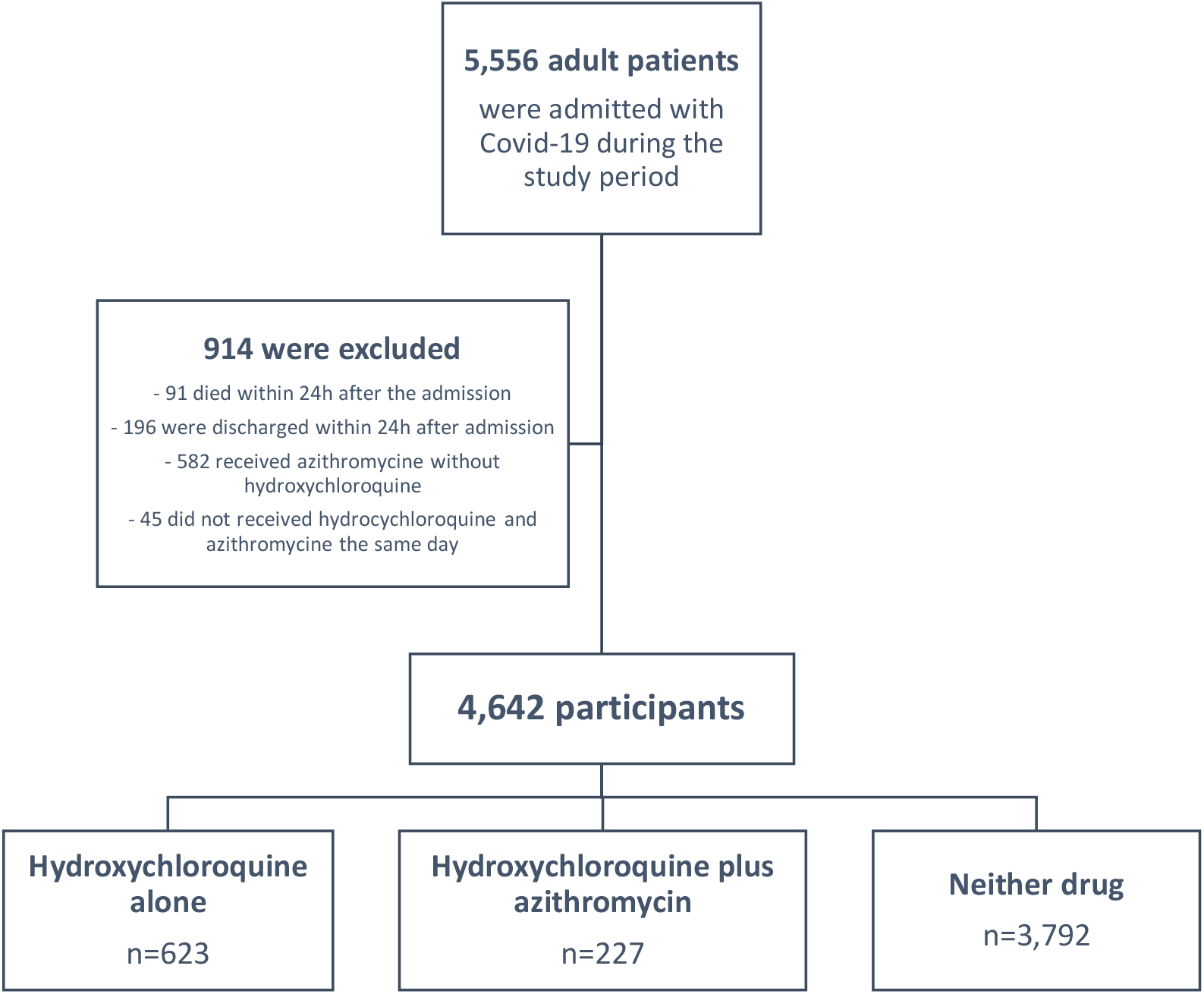
Flow chart of the study population.

### Descriptive results

The main characteristics of the study population are summarized in **Table 1**. Patients receiving ‘HCQ alone’ or ‘HCQ plus AZI’ were more likely younger, males, current smokers and overall presented with slightly more co-morbidities (obesity, diabetes, any chronic pulmonary diseases, liver diseases) than the ‘Neither drug’ group, while no major difference was apparent in biological parameters.

**Table 1.**
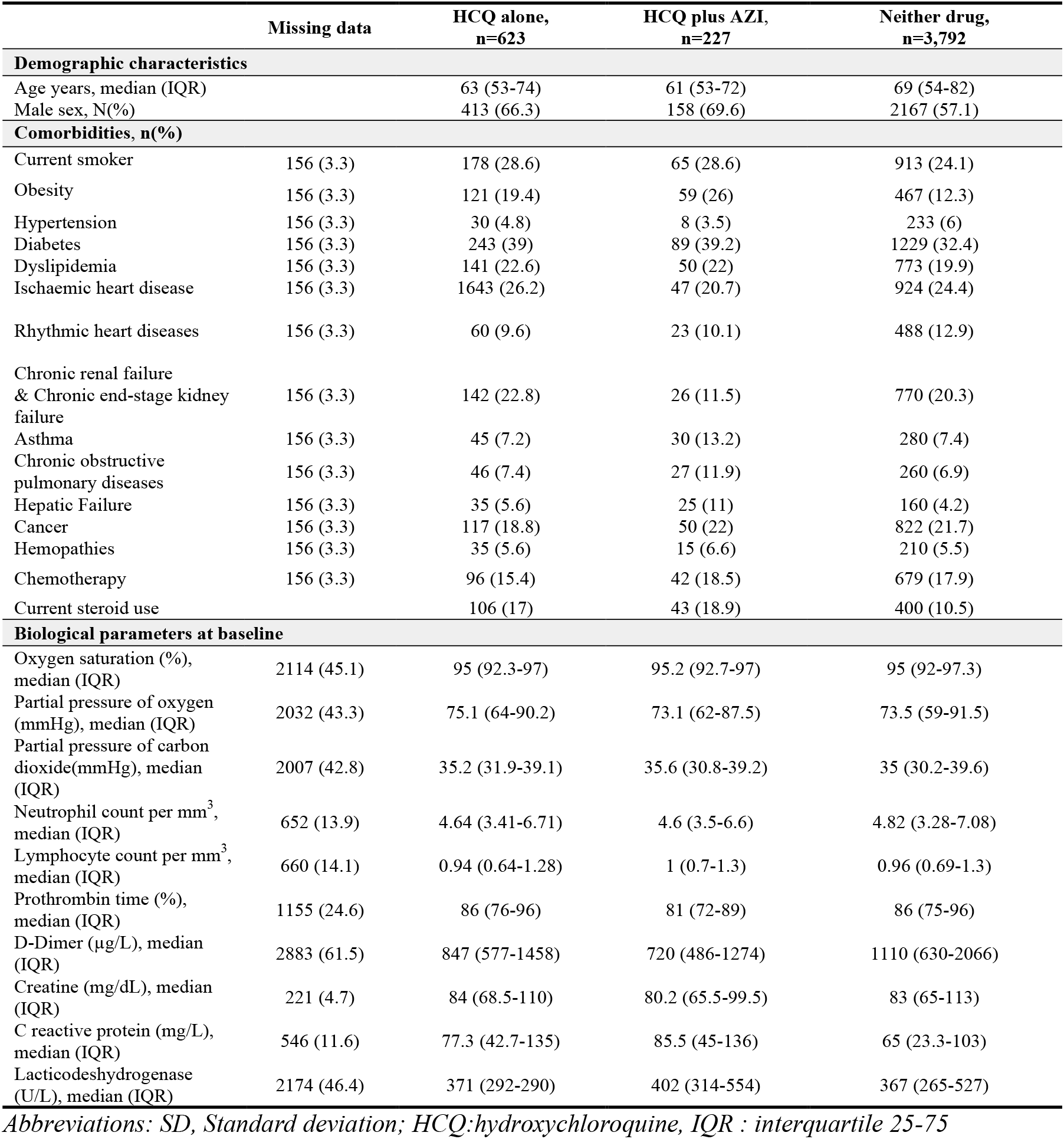
Patients characteristics by treatment group

**Table 2** shows unadjusted clinical outcomes by treatment group. Raw 28-day mortality rates statistically differed between the three groups (number of deaths: 111 (17.8%), 54 (23.8%) and 830 (21.9%) for ‘HCQ alone’, ‘HCQ plus AZI’ and ‘Neither drug’ groups, respectively; p<0.001). Of the 4,642 patients, 777 (16.7%) were transferred to ICU within 24h after the admission, more markedly so in the ‘HCQ plus AZI’ group (27.3%; p<0.001). Groups main characteristics stratified by early ICU transfer are summarized in **Supplemental Tables S2 and S3**. Discharge rates at day 28 significantly varied across groups, ranging from 39.7% (neither drug) to 56.3% (HCQ alone; p<0.001), with corresponding lengths of stay among patients who lived ranging in median (IQR) from 9.8 [6.9-17.6](HCQ+AZI) to 10.9 [4.7-31.9](neither drug; p=0.974) and in mean±SD from 14.7±12 (HCQ+AZI) to 17.6±15.7 (neither drug, p<0.001).

**Table 2.**
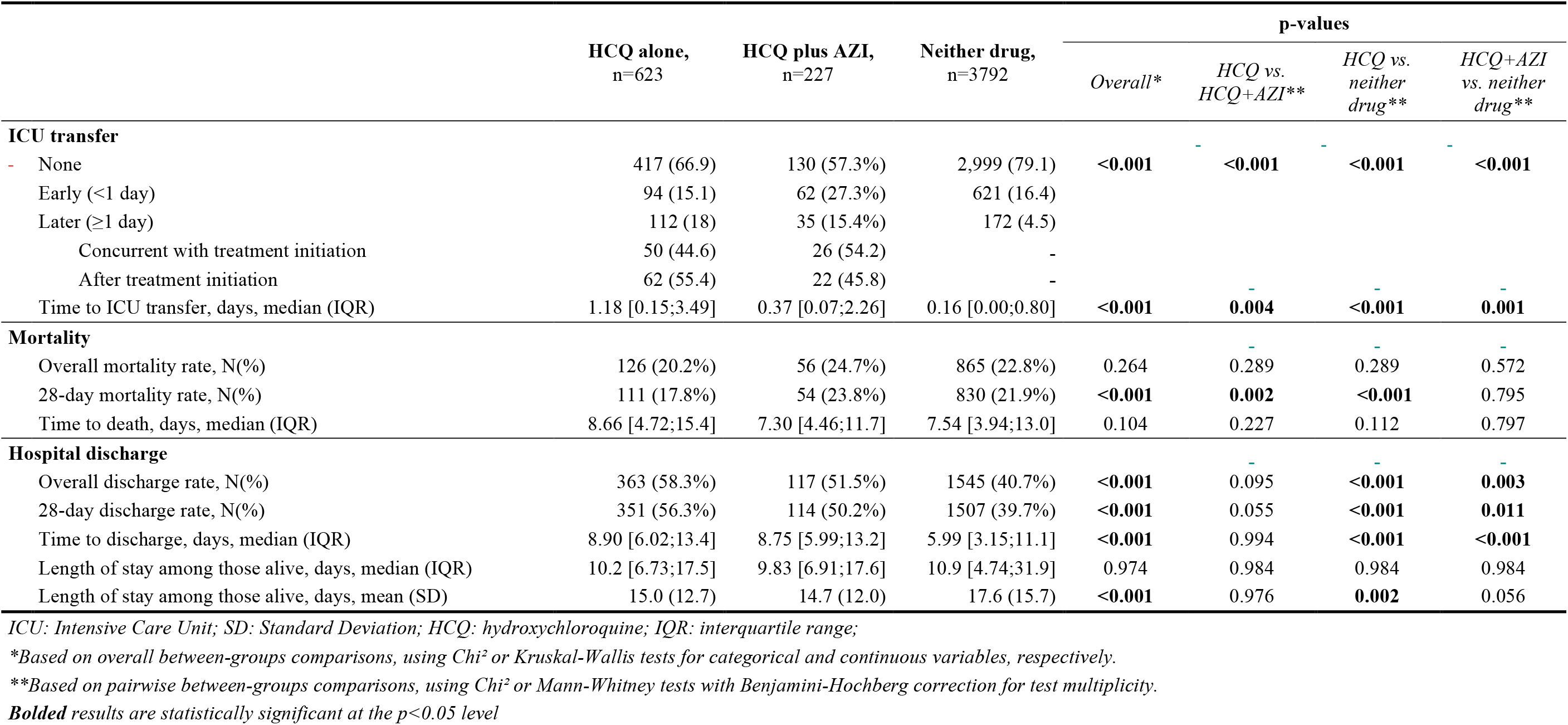
Unadjusted clinical outcomes by treatment group

### Multivariable analyses

Results from competing risks multivariable analyses for 28-day mortality and hospital discharge are displayed in **Table 3**, showing both raw unadjusted estimates for the average treatment effect of ‘HCQ alone ‘or ‘HCQ plus AZI’, and AIPTW results from double robust estimation accounting for confounders for the outcome and the treatment allocation.

**Table 3.** Primary and secondary outcomes according to study population and treatment group

|  |  | HCQ alone<br>vs.<br>neither drug |  | HCQ plus AZI<br>vs.<br>neither drug |  |
| --- | --- | --- | --- | --- | --- |
|  |  | Raw Estimate (95%CI) | AIPTW Estimate*<br>(95%CI) | Raw Estimate (95%CI) | AIPTW Estimate*<br>(95%CI) |
| *Whole population* |  |  |  |  |  |
| **28-day mortality** | Difference in average treatment effect | **-5.59% (-9.12 to -2.05)** | +1.24% (-5.63 to 8.12) | +1.19% (-5.02 to 7.39) | +9.83% (-0.51 to 20.17) |
|  | Ratio in average treatment effect | **0.78 (0.64 to 0.91)** | 1.05 (0.77 to 1.33) | 1.05 (0.80 to 1.30) | 1.40 (0.98 to 1.81) |
| **28-day hospital discharge** | Difference in average treatment effect | **+16.4% (12.0 to 20.8)** | **+11.1% (3.30 to 18.9)** | **+12.2% (5.08 to 19.4)** | +0.16% (-9.97 to 10.29) |
|  | Ratio in average treatment effect | **1.37 (1.27 to 1.48)** | **1.25 (1.07 to 1.42)** | **1.28 (1.11 to 1.44)** | 1.00 (0.77 to 1.23) |
| *Population who were transferred to ICU within the first 24h* |  |  |  |  |  |
| **28-day mortality** | Difference in average treatment effect | -5.25% (-15.28 to 4.77) | -7.83% (-24.97 to 9.32) | -0.39% (-13.18 to 12.40) | +6.20% (-7.05 to 19.45) |
|  | Ratio in average treatment effect | 0.84 (0.53 to 1.14) | 0.75 (0.20 to 1.30) | 0.99 (0.59 to 1.38) | 1.20 (0.77 to 1.63) |
| **28-day hospital discharge** | Difference in average treatment effect | **+14.63% (3.43 to 25.84)** | +0.76% (-17.55 to 19.08) | **+18.2% (4.51 to 32.0)** | +2.74% (-7.19 to 12.67) |
|  | Ratio in average treatment effect | **1.51 (1.09 to 1.93)** | 1.03 (0.42 to 1.64) | **1.64 (1.13 to 2.15)** | 1.09 (0.76 to 1.42) |
| *Population who were not transferred to ICU within the first 24h* |  |  |  |  |  |
| **28-day mortality** | Difference in average treatment effect | **-5.32% (-9.08 to -1.56)** | +3.01% (-4.14 to 10.15) | +0.48% (-6.55 to 7.51) | +9.54% (-4.20 to 23.27) |
|  | Ratio in average treatment effect | **0.77 (0.62 to 0.93)** | 1.13 (0.82 to 1.44) | 1.02 (0.72 to 1.32) | 1.41 (0.82 to 2.00) |
| **28-day hospital discharge** | Difference in average treatment effect | **+16.3% (11.6 to 21.1)** | +7.42% (-0.72 to 15.55) | **+12.7% (4.43 to 20.9)** | +0.34% (-12.28 to 12.96) |
|  | Ratio in average treatment effect | **1.35 (1.24 to 1.45)** | 1.16 (0.98 to 1.33) | **1.27 (1.09 to 1.45)** | 1.01 (0.74 to 1.28) |

\*AIPTW: Augmented inverse probability of treatment weight estimator for doubly robust inference of the average treatment effect conditional on baseline covariates, derived from cause specific Cox proportional hazards modeling; Factorial Analysis for Mixed Data model was used for handling missing data; 95%CI: 95% confidence interval
Baseline covariables considered for adjustment were sex, age, current smoker, diabetes, obesity, hypertension, dyslipidemia, Ischaemic heart disease, rhythmic heart diseases, Chronic renal failure & Chronic end-stage kidney failure, any chronic lung disease, hepatic failure, cancer, hemopathies, chemotherapy, current steroid use, oxygen saturation, partial pressure of oxygen, $paCO_2$ , lymphocytes, neutrophils, D-Dimer, Creatine, C Reactive protein, Dehydrogenase lactate, prothrombin time
**Bolded** results are statistically significant at the $p < 0.05$ level

In the whole population, the raw difference in average treatment effect for ‘HCQ’ versus ‘neither drug’ comparison was −5.59% (−9.12 to −2.05); in other words, there was a 5.59% absolute reduction in the 28-day mortality rate in the ‘HCQ alone’ group when compared to patients in the ‘Neither drug’ group, with a corresponding ratio in ATE of 0.78 (0.64 to 0.91). After accounting for confounding, no significant difference was observed between the ‘HCQ alone’ and ‘Neither drug’ groups: AIPTW difference in ATE of +1.24% (−5.63 to 8.12), ratio in ATE of 1.05 (0.77 to 1.33; p=0.723). Regarding 28-day discharge rates, a statistically significant difference was found for the ratio in AIPTW ATE in favor of ‘HCQ alone’ (1.25 [1.07 to 1.42; p=0.006]). For the ‘HCQ plus AZI’ vs. ‘Neither drug’ comparison, a trend was found towards higher mortality rates in the former group, though not reaching stistical significance (difference in AIPTW ATE +9.83% [−0.51 to 20.17], ratio in ATE 1.40 [0.98 to 1.81]; p=0.062).

In subgroup analyses stratified by early ICU tranfer or not, results were close to those described above, with no statistically significant difference observed in AIPTW ATE between ‘HCQ alone’ and ‘Neither drug’ groups for the 28-day mortality, a trend towards higher discharge rates for the ‘HCQ alone’ group in the not-early ICU transfer subpopulation (ratio in AIPTW ATE 1.13 [0.98 to 1.33]; p=0.075), and apparent worse mortality rates in the ‘HCQ plus AZI’ group but without reaching statistical significance.

### Sensitivity analyses

Results from double robust analyses considering 28-day mortality as a binary endpoint analyzed at a fixed timepoint are shown in **Supplemental Table S4**, with corresponding balance plots of covariates before and after IPT-weighting depicted in **Supplemental Figure S4**. These analyses confirmed those obtained for ‘HCQ alone’ group from the main analysis with no statistically significant difference in mortality, but found no excess of mortality for the ‘HCQ plus AZI’ group compared with ‘Neither drug’ group.

Results from multivariable analyses using conventional adjustment and/or IPT weighting are shown in **Figure 2** for the cause specific Cox proportional hazards models. Using these approaches, results essentially confirmed those obtained from double robust estimates, illustrating for the ‘HCQ alone’ vs. ‘Neither drug’ comparison (**Figure 2A**) the influence of confounders on estimations as indicated by the progressive alignment of death incidence curves according to treatment after adjustement and/or IPT weighting, and the persistence of statistically significant differences in discharge rates after adjustment, but not when using IPT weighting (p=0.073) or combining both approaches (p=0.116). Regarding the ‘HCQ plus AZI’ vs. ‘Neither drug’ comparison (**Figure 2B**), a trend for a statistically significant difference was found for mortality after multiple adjustment and IPT weighting (HR=1.53; p=0.057), but not for discharge (HR=0.98; p=0.923). Results from Fine-Gray models identified similar but not significant trends for the excess risk of mortality in the ‘HCQ plus AZI’ group. (**Supplemental Figures S2 and S3)**.

**Figure 2.**
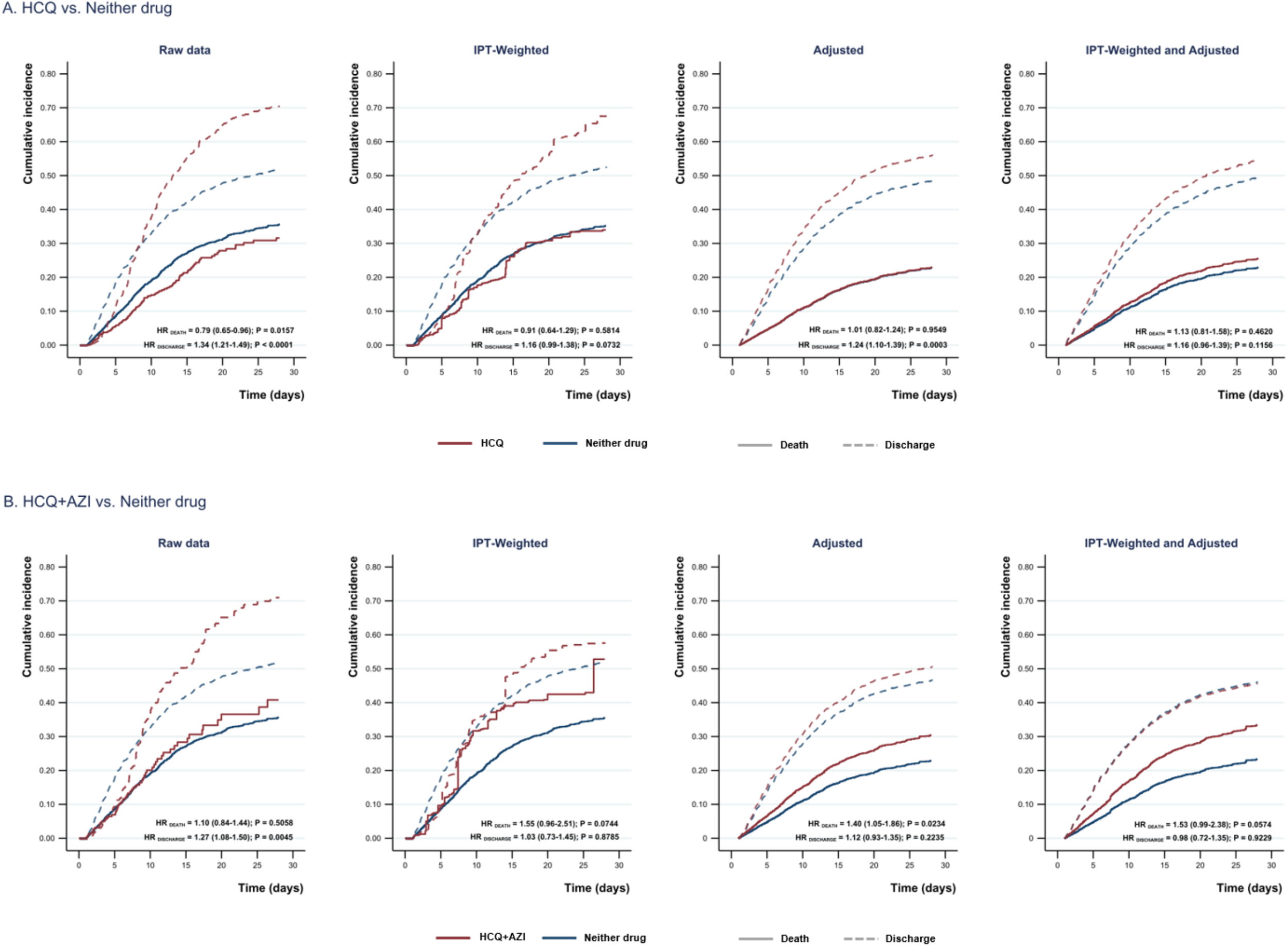
Death and discharge cumulative incidence curves: results from cause specific Cox competing risks analysis. Panel A - HCQ vs. Neither drug; Panel B - HCQ+AZI vs. Neither drug; Adjusted results are based on the adjustment strategy for the outcome detailed in Figure S1

## Discussion

Using a large non-selected population of 4,642 hospitalized for COVID-19 infection in 39 hospitals in France, we found no evidence for efficacy of HCQ or HCQ combined with AZI on 28-day mortality. Our findings suggest that patients treated by association of HCQ and AZI are at greater risk of mortality compared with the ‘Neither drug’ group. Interestingly, significantly higher rates of discharge home were observed in patients treated by HCQ when using competing risks survival analyses, a finding whose statistical significance persisted after multivariable adjustment and propensity-score weighting. These results were found to be robust to a variety of methodological approaches conducted regarding missing data handling and causal inference modeling to properly account for potential confounders.

The absence of difference on mortality between hospitalized patients receiving HCQ and those receiving neither drug is consistent with previous observational studies led in hospitalized patients.^8–11^ Similarly to two recent studies with a >1000 sample size, patients from the present report had a median age around 65 years, with substantial differences at baseline between patients being prescribed HCQ and those being not, including higher prevalence in the former group of co-morbidities and risk factors such as obesity, diabetes, hypertension,^10^ chronic lung and cardiovascular diseases.^11^ However, patients with either drug in our study were remarkably younger (63 [HCQ], 61 [HCQ+AZI], 69 [neither drug]) and fared significantly better in raw analyses, in contrast with findings from the two previous studies where worse raw outcomes were observed in the groups receiving HCQ. Yet in all studies, including ours, no statistically significant difference was eventually found after accounting for confounding by multivariable analyses potentially combined with propensity score weighting, matching or adjustment methods. Finally, it should be stressed out that findings from these reports pertain to hospitalized patients and do not provide information on the efficacy of either drug when administered in earlier forms of the disease.

Regarding the association of HCQ and AZI, our findings indicate a trend towards higher risk of death for the ‘HCQ plus AZI’ group compared with the ‘Neither drug’ group (ratio in ATE at 1.40 [0.98 to 1.81]). This finding is consistent with the results from Rosenberg et al study (adjusted HR for mortality 1.35 [0.76 to 2.40]). Because of a limited sample size for this subgroup in our study, our results should be taken with caution. However, among possible hypotheses that could be discussed, an increased risk of serious adverse events such as arrhythmia has been advocated in several studies.^11,28^ Among 90 consecutively included patients receiving HCQ for Covid-19 infection in Israel, Mercuro et al detected change in QTc in 21 patients (23%), and more evidently so in the ‘HCQ plus AZI’ subgroup.^28^ HCQ is already known to inhibit voltage-gated sodium and potassium channels, prolonging the QT interval and increasing the risk of torsades de pointes, syncope and sudden cardiac death.^29^ Azithromycin has also been implicated in QTc prolongation and proarrhythmic events.^29^ In addition to HCQ and AZI interaction, patients hospitalized with severe COVID-19 pneumonia are also at risk to present clinical characteristics leading to QT prolongation such as hypokalemia or congestive heart failure.

One original finding of the present paper lays in the assessment of discharge and in-hospital death in a competing events survival analysis framework. This analysis allowed us to investigate both events separately, to provide insights beyond that provided by mortality rates alone. In our study, we identified increased discharge rates at day-28 in the HCQ group, corresponding to a ratio in average treatment effect of 1.28 in favor of HCQ, with corresponding predicted day-28 discharge rates of 56% [HCQ] vs. 45% [neither drug] in the whole study population. To our knowledge, this is the first report from a large observational study specifically addressing this issue in addition to mortality and ICU-related outcomes,^8–11^ whereas a similar endpoint was used for remdesivir randomized controlled trials for COVID 19 infection as a primary outcome.^30^ While mortality remains the ultimate clinical endpoint, assessing discharge rates and determinants may inform decision making for optimizing organization of inpatient services in the context of a pandemic.^31^ Our results could be supported by in vitro and in vivo studies which established a decrease of the viral load in patients receiving HCQ and HCQ plus AZI.^3,6^ Causal interpretation should here again remain cautious, but should such results be replicated and validated in randomized studies, it would provide useful information to draw a balanced assessment of the risks and benefits associated with HCQ-based regimens. From a methodological standpoint, the concomitant examination of in-hospital mortality and discharge and length of stay through survival analyses require to account for the effect of competing risks to avoid overestimation of the probability of the events of interest.^32^ With that regard, we used two approaches to competing risks, primarily using cause specific Cox models for causal inference, and further confirming our results by using the Fine-Gray approach with IPT weighting and multivariable adjustment.

Among the strengths of our study is the use of advanced causal inference approaches both considering time-to-event survival analyses and binary endpoints at a single timepoint. Because inappropriately accounting for confounders can drastically modify results, we performed several sensitivity analyses to check the stability of our results under varying approaches, including so called double robust estimations relying on both multivariable modeling of the outcome and of the treatment (including propensity score-based approaches), which offer better robustness to model misspecification, and use of varying missing data imputation techniques confirming the stability of our findings. Other strengths of this work include the assessment of a large, representative sample study population in the Greater Paris area from 39 hospitals. Study’s limitations include the absence of direct, clinical information on regimen duration and dosages, and respiratory parameters of COVID-19 infection, including oxygen requirement, non-invasive or mechanical ventilation, which are potential confounders. However, we used biological parameters proxy to assess the severity of the COVID-19 infection including creatine, lymphocyte count and inflammatory markers (D-Dimer and C-Reactive protein) well known to be associated with severity of COVID-19^21^Yet, causal interpretation of our findings relying on retrospective evaluation of medical records should remain cautious considering the observational nature of the study design.

## Conclusion

Using a large non-selected population of inpatients hospitalized for COVID-19 infection in 39 hospitals in France and robust methodological approaches, we found neither evidence for reduced or excess risk of 28-day mortality with the use of HCQ alone. Our findings suggest a possible higher risk of death for patients receiving HCQ combined with AZI. Significantly higher rates of discharge home were observed in patients treated by HCQ, a novel finding warranting further confirmation in replicative studies.

## Data Availability

The data are available on request after publication.

## Acknowledgments

We thank Susan Wilkinson for editorial assistance.

Data used in preparation of this article were obtained from the AP-HP Covid CDR Initiative database. A complete listing of ECAI members can be found at (https://eds.aphp.fr/covid-19).

## Funding

This research did not receive any specific grant from funding agencies in the public, commercial, or not-for-profit sectors.

## Competing interests

All authors have completed the ICMJE uniform disclosure form at www.icmje.org/coi_disclosure.pdf and declare: Armand Mekontso Dessap reports research grants from Fischer Paykel, Baxter, Philips, Ferring and GSK; participation to advisory board for Air liquid, Baxter, and Amomed; lectures for Getingu and Addmedica, all outside the scope of the submitted work; Etienne Audureau reports (2019) consulting for consulting GBT and HEMANEXT (both protocol design of an unrelated work in hematology). The other authors : no support from any organisation for the submitted work; no financial relationships with any organisations that might have an interest in the submitted work in the previous three years; no other relationships or activities that could appear to have influenced the submitted work.

## Ethical approval

This study was approved by the French data protection agency Commission Nationale de l’Informatique et des Libertés (regulatory decision DE-2017-013), IRB00011591.

## Data sharing

The data are available on request.

## Transparency

The manuscript’s guarantors (ES) affirm that this manuscript is an honest, accurate and transparent account of the study being reported; that no important aspects of the study have been omitted; and that any discrepancies from the study as planned (and, if relevant, registered) have been explained.

## Contributors

Contributors: ES, JJ, GL, IM, MB, AG, GV, ML and EA conceived and designed the experiments. JJ, IM, ML and EA performed the experiments. ES, JJ, GL, IM, MB, AG, GV, ML and EA analysed the data. ES, JJ, PW, EC, ML, AMD and EA interpreted the results. ES and EA wrote the first draft of the manuscript. All the authors contributed to the writing of the manuscript. All the authors agreed with the results and conclusions of the manuscript. All authors have read, and confirm that they meet, ICMJE criteria for authorship. All authors had full access to all of the data (including statistical reports and tables) in the study and can take responsibility for the integrity of the data and the accuracy of the data analysis. ES is the guarantor

## Notes

### Author Declarations

This study was approved by the French data protection agency Commission Nationale de l Informatique et des Libertes (regulatory decision DE-2017-013), IRB00011591.

